# Seasonality of drug-related hypoglycemia in Japan: A study using spontaneous reporting system data

**DOI:** 10.64898/2026.01.08.26343703

**Authors:** Yuka Sakazaki, Yuki Kondo, Yoichi Ishitsuka

## Abstract

Hypoglycemia is a common issue in diabetes pharmacotherapy, with seasonal variations potentially influencing its occurrence. It has been reported that blood glucose levels exhibit seasonal variation, suggesting that seasonal factors may influence the occurrence of hypoglycemia. In this study, we examined the seasonal patterns of drug-related hypoglycemia according to causative drug categories using the Japanese Adverse Drug Event Report database. We assessed a total of 545,012 adverse event reports submitted between December 2004 and November 2024, among which 5332 cases of drug-related hypoglycemia were identified. The results indicated a consistent increase in hypoglycemia reports during the winter months, with the lowest percentage observed in August (0.78%), followed by a gradual rise, reaching a peak in February (1.13%). Diabetic medications, including sulfonylureas, alpha-glucosidase inhibitors, and thiazolidinediones, showed an increased frequency of hypoglycemia signals in winter. Some non-diabetic medications, such as fluoroquinolones and angiotensin II receptor blockers, exhibited similar seasonal trends. These findings suggest that seasonal factors may warrant consideration in the prevention of drug-related hypoglycemia.

## Introduction

The occurrence of hypoglycemia remains an important challenge in the pharmacotherapy of diabetes. Although most episodes of hypoglycemia are transient, severe cases can require hospitalization and may even result in death, leading to substantial medical costs[1]. Additionally, severe hypoglycemia has been associated with a range of adverse outcomes, including subsequent cardiovascular disease[2], dementia[3,4], traffic accidents[5] and an increased risk of fractures due to falls[6], underscoring the urgent need to prevent hypoglycemia. Several risk factors for hypoglycemia have been identified, such as age, serious comorbidities (e.g., sepsis, renal dysfunction, liver dysfunction), and individual patient characteristics, including the duration of diabetes mellitus[7]. Furthermore, it has been suggested that the incidence of hypoglycemia in patients with diabetes is higher during the winter months, indicating potential seasonal variation in hypoglycemia occurrence. In these previous studies investigating seasonal variation, most reported cases involved the use of sulfonylureas (SUs)[8,9]. However, it remains unclear whether similar seasonal patterns of hypoglycemia occur with other classes of antidiabetic drugs. Furthermore, several non-antidiabetic medications, such as quinolone antibiotics[10,11] and antidepressants[12], have also been reported to induce hypoglycemia, but little is known about the seasonality of hypoglycemia associated with these drugs[13]. The aim of this study was to examine the seasonality of drug-related hypoglycemia according to causative drug categories using the Japanese database of spontaneous adverse event reports.

## Materials and Methods

### Study population

The Japanese Adverse Drug Event Report (JADER) database comprises adverse event reports submitted to the Pharmaceuticals and Medical Devices Agency. The database includes cases reported by medical institutions and pharmaceutical companies. There are four data tables in this database: DEMO (e.g., sex and age), DRUG (e.g., drug name and route of administration), REAC (e.g., adverse events and dates of adverse events), and HIST (e.g., comorbidities). In the present study, we included 545,012 patients with adverse events reported between December 2004 and November 2024, who had no missing information on patient sex or age; the database was accessed on May 3, 2025.

### Definitions

In JADER, adverse events and comorbidities are coded using the preferred terms (PTs) in the Medical Dictionary for Regulatory Activities (MedDRA). In this study, the Japanese version of MedDRA (MedDRA/J) ver. 26.0 was used. Hypoglycemia events were identified using the narrow definition of hypoglycemia [20000226] in the Standardized MedDRA Query. If a hypoglycemia event was reported more than once during the study period in the same patient, the earliest onset date was used. To determine whether a case had pre-existing diabetes whenever possible, cases with a narrow definition of hyperglycemia/diabetes onset [20000041] in the Standardized MedDRA Query or PTs related to diabetic complications (e.g., diabetic nephropathy and diabetic retinopathy) in the HIST table were considered to have pre-existing diabetes. PTs used for identification of hypoglycemia and diabetes events are shown in S1 Table.

Drugs were classified according to medicinal class using the Anatomical Therapeutic Chemical classification. We excluded from the analysis any drugs reported in the JADER database for which the ingredient name was unknown. We conducted a seasonal analysis for each of the 15 drug categories most frequently reported as causative agents of hypoglycemia. Drug categories with a limited number of reports were excluded to ensure the stability of seasonal estimates. The corresponding number of reports is provided in S2 Table.

Seasons were classified as spring (March–May), summer (June–September), autumn (October–November) and winter (December–February). Based on recent climatological data in Japan, September continues to exhibit temperature patterns similar to summer months and was therefore grouped with summer in the seasonal analysis.

### Statistical analysis

To assess the seasonality of hypoglycemia, the percentage of hypoglycemia reported per month, and the reporting odds ratio (ROR) and 95% confidence interval (CI) for reports of hypoglycemia were calculated. The ROR was adjusted for age, sex, and reporting year using logistic regression. In calculating the ROR, August—which had the lowest reported rate of hypoglycemia—was used as the control month for monthly assessment; summer—which had the lowest reported rate of hypoglycemia—was used as the target season for seasonal assessment. The formula used to calculate the ROR is:

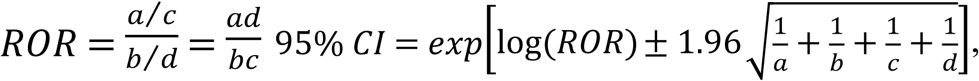

where *a* refers to cases identified as hypoglycemia in that month (season); *b* refers to cases identified as hypoglycemia in the control month (season); *c* refers to adverse event cases other than hypoglycemia in that month (season); and *d* refers to adverse event cases other than hypoglycemia in the control month (season). If the lower limit of 95% CI for the ROR was greater than 1, this indicated a signal of increased hypoglycemia for that month. The statistical analysis was performed using JMP^®^ Student Edition 18.2 software (SAS Institute Inc., Cary, NC, USA).

## Results

### Patient characteristics

The characteristics of hypoglycemia cases are shown in Table 1. A total of 5332 cases of drug-related hypoglycemia were analyzed during this period. Among all cases, most were male (51.9%), and approximately 77.7% were aged 60 years or older. Pre-existing diabetes was present in 48.4% of cases (n = 2584). SUs were the most frequently reported drugs related to hypoglycemia (n = 1696, 31.8%).

**Table 1.** Characteristics of patients with drug-related hypoglycemia.

| Variable |  | n (%) |
| --- | --- | --- |
| Sex | Male | 2765 (51.9) |
|  | Female | 2567 (48.1) |
| Age | <10s | 213 (4.0) |
|  | 10s | 54 (1.0) |
|  | 20s | 75 (1.6) |
|  | 30s | 142 (2.7) |
|  | 40s | 256 (4.8) |
|  | 50s | 438 (8.2) |
|  | 60s | 983 (18.4) |
|  | 70s | 1508 (28.3) |
|  | 80s | 1381 (25.9) |
|  | 90s | 268 (5.0) |
|  | 100s | 3 (0.1) |
| Drugs used for diabetes | Sulfonylureas | 1696 (31.8) |
|  | Insulins and analogs for injection, fast-acting | 1359 (25.5) |
|  | Insulins and analogs for injection, long-acting | 1154 (21.6) |
|  | Dipeptidyl peptidase 4 inhibitors | 2090 (20.4) |
|  | Alpha-glucosidase inhibitors | 790 (14.8) |
|  | Biguanides | 619 (11.6) |
|  | Other blood glucose-lowering drugs, excluding insulins | 311 (5.8) |
|  | Thiazolidinediones | 370 (6.9) |
|  | Sodium-glucose cotransporter 2 inhibitors | 183 (3.4) |
|  | Glucagon-like peptide-1 analogs | 138 (2.6) |
| Drugs used for non-diabetes | Fluoroquinolones | 481 (9.0) |
|  | Other antiarrhythmics, class I and III | 288 (5.4) |
|  | Angiotensin II receptor blockers, plain | 1105 (20.7) |
|  | Other antipsychotics | 156 (2.9) |
|  | Diazepines, oxazepines, thiazepines, and oxepines | 126 (2.4) |

### Seasonality of drug-related hypoglycemia reported in JADER for all drugs

The seasonality of drug-related hypoglycemia reported in JADER for all drugs is shown in Fig 1. The percentage of drug-related hypoglycemia was lowest in August (0.78%) and gradually increased toward February (1.13%). Similarly, the adjusted RORs increased gradually during the winter months: December (adjusted ROR: 1.33, 95% CI: 1.16–1.53), January (adjusted ROR: 1.28, 95% CI: 1.12–1.47), and February (adjusted ROR: 1.37, 95% CI: 1.19–1.57).

**Fig 1.**
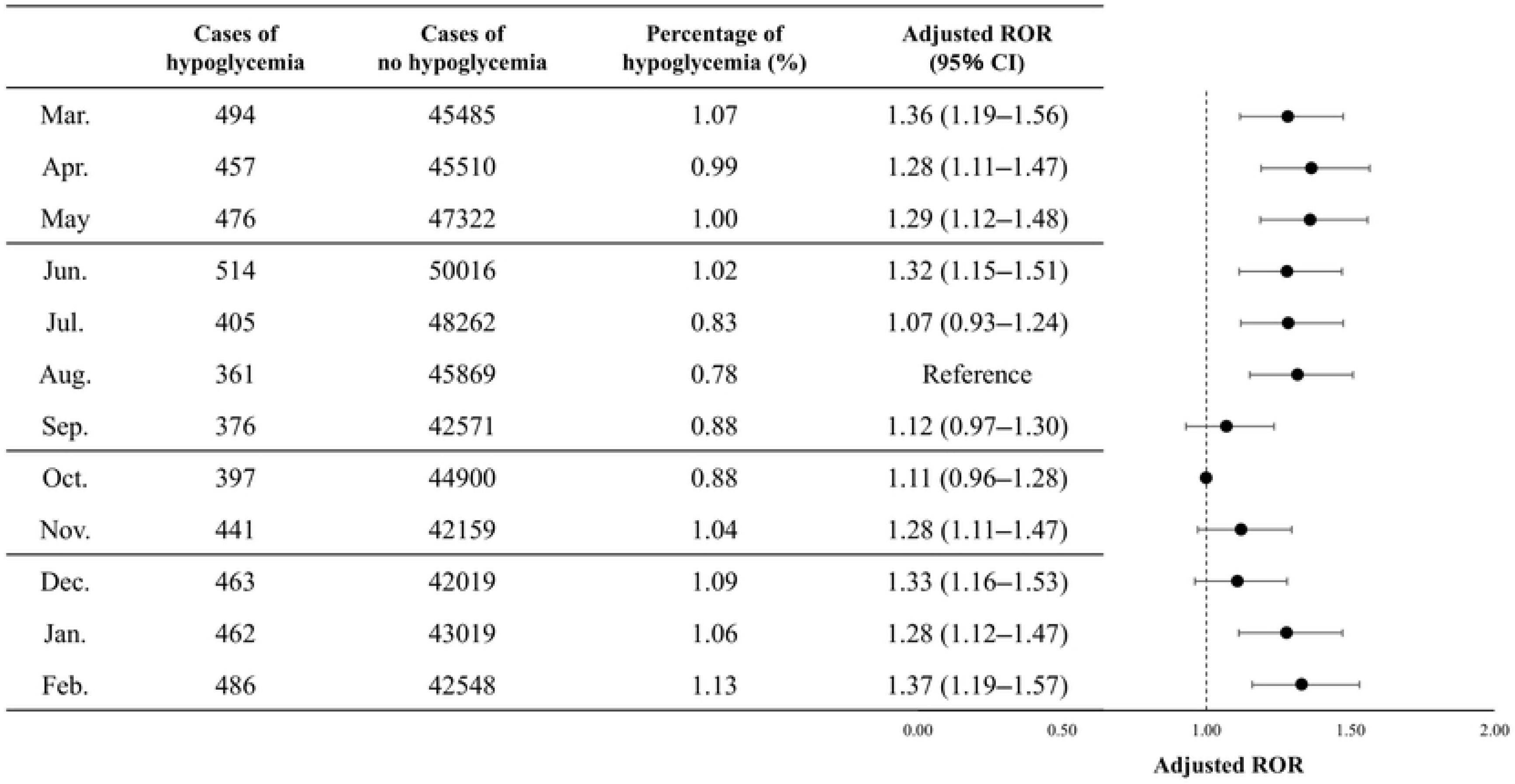
Seasonality of drug-related hypoglycemia in all drugs reported in the Japanese Adverse Drug Event Report (JADER) database.

The percentage of drug-related hypoglycemia reports was consistently lower in the warmer months from July to September and higher in the colder months from December to January, regardless of the year of occurrence (Fig 2).

**Fig 2.**
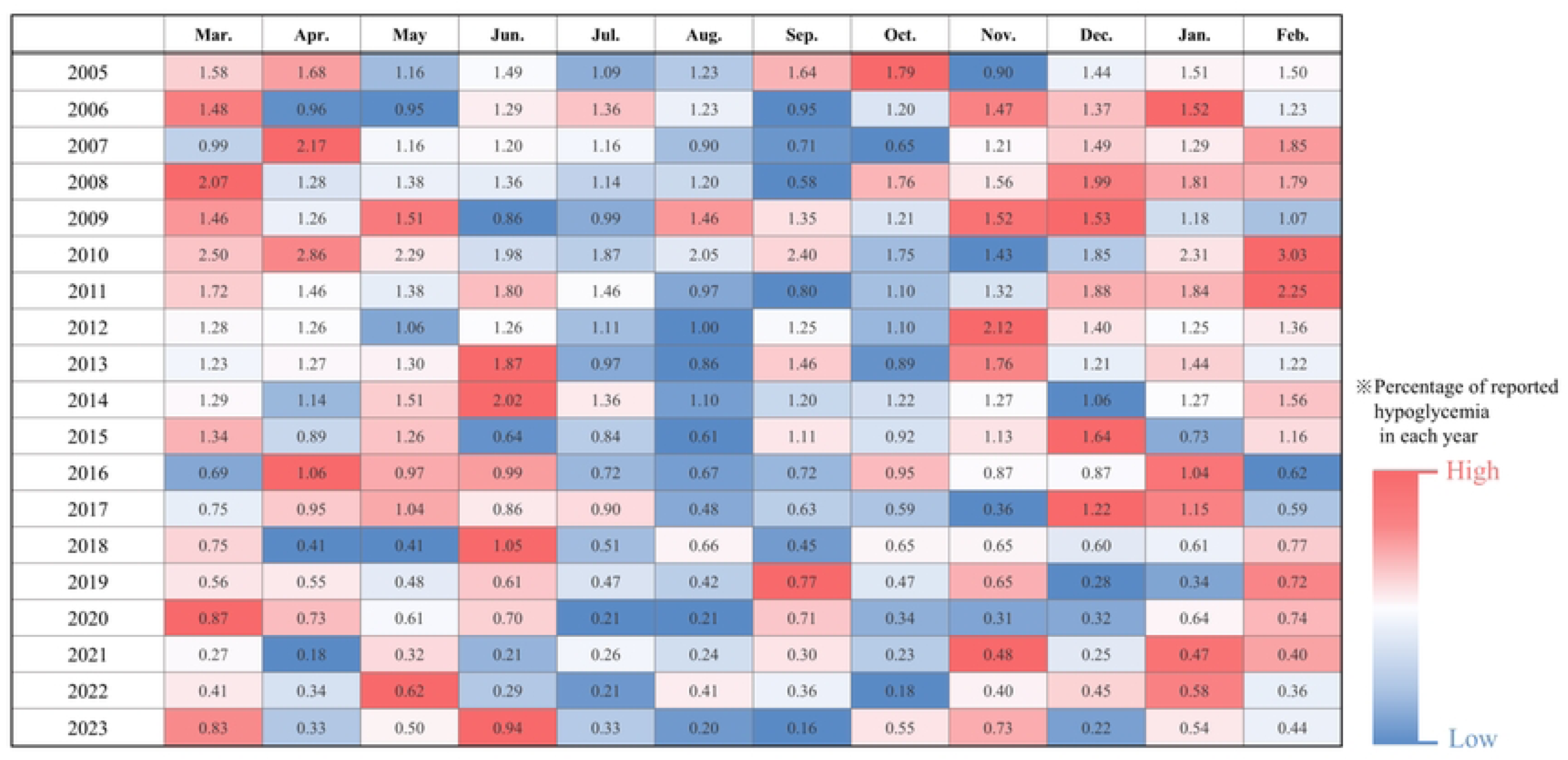
Monthly trends in the percentage of reported drug-related hypoglycemia among adverse events (2005–2023).

Values represent the percentage of hypoglycemia reports relative to total reported adverse events for each month. The color gradient indicates the relative proportion within each year, where red denotes a higher percentage and blue denotes a lower percentage.

### Seasonality of drug-related hypoglycemia related to diabetes medications

Table 2 presents the results of seasonality analysis for diabetic medications, categorized by drug class. Medications that exhibited significantly higher adjusted RORs and 95% CIs for hypoglycemia in winter compared with summer included SUs (adjusted ROR: 1.38, 95% CI: 1.20–1.59, *P*<0.001), alpha-glucosidase inhibitors (adjusted ROR: 1.38, 95% CI: 1.14–1.68, *P*=0.001), and thiazolidinediones (adjusted ROR: 1.50, 95% CI: 1.13–1.98, *P*=0.005). Additionally, alpha-glucosidase inhibitors and thiazolidinediones were more frequently used in combination with SUs, as compared with other diabetic medications (alpha-glucosidase inhibitors: 38.9%, thiazolidinediones: 47.0%).Seasonality of drug-related hypoglycemia related to non-diabetes medications Table 3 presents the results of seasonality analysis for non-diabetic medications by drug class. Medications with significantly higher adjusted RORs and 95% CIs for hypoglycemia in winter compared with summer included systemic anti-infectives, such as fluoroquinolones (adjusted ROR: 1.48, 95% CI: 1.16–1.90, *P*=0.002); cardiovascular agents, specifically angiotensin II receptor blockers (plain) (adjusted ROR: 1.28, 95% CI: 1.09–1.49, *P*=0.003); neuroleptic drugs, including other antipsychotics (adjusted ROR: 1.89, 95% CI: 1.24–2.87, *P*=0.003); and diazepines/oxazepines/thiazepines/oxepines (adjusted ROR: 1.67, 95% CI: 1.05–2.64, *P*=0.029). Among these, angiotensin II receptor blockers (plain) had the highest rate of concomitant use with diabetic medications (27.1%).

**Table 2.**
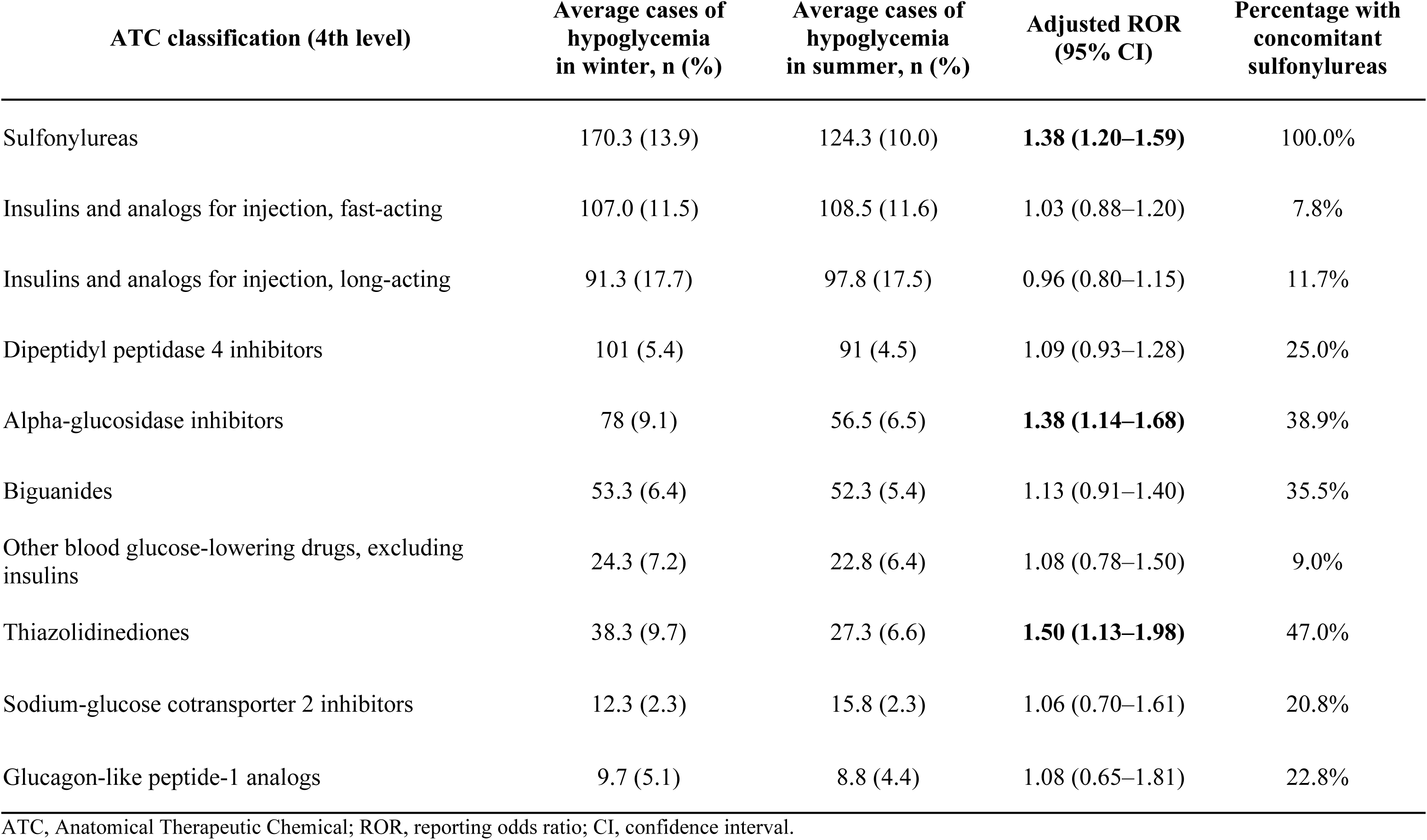
Seasonality of drug-related hypoglycemia for diabetes drugs.

| ATC classification (4th level) | Average cases of hypoglycemia in winter, n (%) | Average cases of hypoglycemia in summer, n (%) | Adjusted ROR (95% CI) | Percentage with concomitant sulfonylureas |
| --- | --- | --- | --- | --- |
| Sulfonylureas | 170.3 (13.9) | 124.3 (10.0) | **1.38 (1.20–1.59)** | 100.0% |
| Insulins and analogs for injection, fast-acting | 107.0 (11.5) | 108.5 (11.6) | 1.03 (0.88–1.20) | 7.8% |
| Insulins and analogs for injection, long-acting | 91.3 (17.7) | 97.8 (17.5) | 0.96 (0.80–1.15) | 11.7% |
| Dipeptidyl peptidase 4 inhibitors | 101 (5.4) | 91 (4.5) | 1.09 (0.93–1.28) | 25.0% |
| Alpha-glucosidase inhibitors | 78 (9.1) | 56.5 (6.5) | **1.38 (1.14–1.68)** | 38.9% |
| Biguanides | 53.3 (6.4) | 52.3 (5.4) | 1.13 (0.91–1.40) | 35.5% |
| Other blood glucose-lowering drugs, excluding insulins | 24.3 (7.2) | 22.8 (6.4) | 1.08 (0.78–1.50) | 9.0% |
| Thiazolidinediones | 38.3 (9.7) | 27.3 (6.6) | **1.50 (1.13–1.98)** | 47.0% |
| Sodium-glucose cotransporter 2 inhibitors | 12.3 (2.3) | 15.8 (2.3) | 1.06 (0.70–1.61) | 20.8% |
| Glucagon-like peptide-1 analogs | 9.7 (5.1) | 8.8 (4.4) | 1.08 (0.65–1.81) | 22.8% |

ATC, Anatomical Therapeutic Chemical; ROR, reporting odds ratio; CI, confidence interval.

**Table 3.** Seasonality of drug-related hypoglycemia for non-diabetic drugs.

|  | ATC classification (4th level) | Average cases of hypoglycemia in winter, n (%) | Average cases of hypoglycemia in summer, n (%) | Adjusted ROR (95% CI) | Percentage with concomitant antidiabetic agents |
| --- | --- | --- | --- | --- | --- |
| Anti-infectives for systemic use | Fluoroquinolones | 49.7 (2.7) | 31.3 (1.7) | **1.48 (1.16–1.90)** | 11.4% |
| Cardiovascular system | Other antiarrhythmics, class I and III | 21.7 (19.1) | 25.8 (21.7) | 0.83 (0.57–1.21) | 12.8% |
|  | Angiotensin II receptor blockers, plain | 106.3 (2.3) | 81.5 (1.7) | **1.28 (1.09–1.49)** | 27.1% |
| Nervous system | Other antipsychotics | 16.7 (1.9) | 10.5 (1.0) | **1.89 (1.24–2.87)** | 2.5% |
|  | Diazepines, oxazepines, thiazepines, and oxepines | 13.7 (1.6) | 8.8 (0.9) | **1.67 (1.05–2.64)** | 9.9% |

ATC, Anatomical Therapeutic Chemical; ROR, reporting odds ratio; CI, confidence interval.

## Discussion

This study demonstrated that both the number and proportion of drug-related hypoglycemia reports tended to increase gradually during the winter months, and this pattern was consistently observed across all years analyzed. Among antidiabetic medications, SUs, alpha-glucosidase inhibitors, and thiazolidinediones showed a signal indicating an increased risk of hypoglycemia in winter compared with summer. Similarly, among non-antidiabetic medications, certain agents—such as fluoroquinolones and angiotensin II receptor blockers—also exhibited a seasonal signal suggestive of increased hypoglycemia during the winter months.

In this study, most cases of hypoglycemia were reported in male individuals, and most cases occurred in those aged 60 years or older (Table 1). The prevalence of type 2 diabetes in Japan is known to be higher among male individuals[14]; previous studies have also reported a higher incidence of hypoglycemia in male populations[15]. Older adults are thought to be at greater risk for severe hypoglycemia because they are less likely to recognize early autonomic symptoms such as sweating, palpitations, and finger tremors[7,15,16]. SUs were the most frequently reported suspected drugs in hypoglycemia cases in this study (Table 1). These agents induce hypoglycemia by directly stimulating insulin secretion from pancreatic β-cells. When potentiated by certain factors, SUs can lead to excessive insulin release, thereby increasing the risk of hypoglycemia more readily than other antidiabetic drugs. As such, SU use is considered an independent risk factor for hypoglycemia[9].

In this study, the number and proportion of drug-related hypoglycemia reports for all drugs in the JADER database tended to increase gradually during the winter months, and this trend was consistently observed across all years examined (Fig 1 and 2). This finding aligns with previous reports suggesting a seasonal increase in hypoglycemia during winter[8,9]. Other studies have indicated that the seasonality of hypoglycemia may vary depending on the presence of pre-existing diabetes or the type of diabetes[13]. Although the type of diabetes could not be determined in our dataset owing to missing or unspecified information, we stratified the analysis according to the presence or absence of diabetes as a comorbidity. In contrast to previous findings, no difference in the seasonal variation of hypoglycemia was observed according to the presence or absence of a diabetes history in this study (S1 Fig).

In addition to SUs, which, among antidiabetic medications, are well known for causing hypoglycemia, signals of increased hypoglycemia in winter were also observed for alpha-glucosidase inhibitors and thiazolidinediones (Table 2). However, these drugs had a higher proportion of concomitant use with SUs compared with other antidiabetic agents, and it is possible that the observed seasonality was influenced by the effects of SUs. Indeed, when we stratified the analysis based on the presence or absence of concomitant SU use, none of the drugs showed a signal of increased hypoglycemia in winter in the subgroup without SU co-administration. By contrast, in the subgroup with concomitant SU use, both alpha-glucosidase inhibitors and thiazolidinediones showed signals of increased hypoglycemia in winter (S3 Table). These findings suggest that further investigation is warranted to clarify the role of concomitant SU use in the observed seasonal patterns.

Among non-diabetes medications, fluoroquinolones, angiotensin II receptor blockers, other antipsychotics, and diazepines/oxazepines/thiazepines/oxepines exhibited signals of increased hypoglycemia during winter, consistent with the overall trend (Table 3). Because fluoroquinolones, other antipsychotics, and diazepines/oxazepines/thiazepines/oxepines were associated with relatively low rates of concomitant diabetes medication use, it is unlikely that the observed seasonality was driven by diabetes treatments. Like SUs, fluoroquinolones have been reported to influence insulin secretion from pancreatic β-cells [17,18]. However, class I antiarrhythmic agents, which have reported to affect insulin secretion, did not show a signal of increased hypoglycemia in winter, leaving it unclear whether this mechanism is directly associated with seasonal variation.

Although hyperglycemia is a well-known adverse effect of antipsychotics and diazepine-related compounds, several case reports have also been published of severe hypoglycemia induced by these same drugs[19–21]. Nonetheless, the mechanism underlying hypoglycemia associated with the above agents remains unclear, and the reason why these showed seasonal variation in this study is also unknown. Angiotensin II receptor blockers are known to exert hypoglycemic effects by improving insulin resistance through AT1 receptor antagonism[22]. In this study, the proportion of cases involving concomitant use of diabetes medications was relatively higher for this drug class than for the others; thus, the influence of diabetes medications on the observed seasonality cannot be excluded. This study has some limitations. First, as a hypothesis-generating study using disproportionality analysis to detect signals, the findings cannot be used to conclude that the risk of drug-associated hypoglycemia increases in winter. Likewise, the absence of a detected signal does not necessarily indicate the absence of an increased risk. Therefore, further studies are needed to validate the hypotheses generated in this study. Second, because we used spontaneous reporting data in this study, not all hypoglycemia cases were captured, and the actual number of cases may have been underestimated. Third, the reporting rate of hypoglycemia may have been relatively low owing to an increase in reports of other adverse events. Although the proportion of hypoglycemia reports among all adverse events may have decreased, the absolute number of drug-related hypoglycemia reports tended to increase during the winter. Moreover, there was no observed decrease in the number of other adverse event reports during the winter season.

A main strength of this study is the use of a large spontaneous adverse event reporting database, which enabled the inclusion of a substantially greater number of drug-related hypoglycemia cases in comparison with previous studies. Furthermore, owing to limited case numbers, earlier research did not investigate the seasonality of hypoglycemia according to causative agents. To our knowledge, this is the first study to examine seasonal variation in hypoglycemia stratified by causative agent, and the first to suggest that the seasonality of hypoglycemia may vary depending on the agent involved.

## Conclusion

The findings of this study indicated that drug-related hypoglycemia tends to increase during the winter months, underscoring the potential importance of seasonal factors in its prevention. In addition, the observed seasonal patterns appeared to differ by causative drug category. Further research is warranted to validate these hypotheses.

## Data Availability

The data underlying the results presented in the study are available from (https://www.pmda.go.jp/safety/info-services/drugs/adr-info/suspected-adr/0003.html).

## Acknowledgment

We thank Analisa Avila, MPH, ELS, of Edanz (https://jp.edanz.com/ac) for editing a draft of this manuscript.

## Reference

1. Curkendall SM, Natoli JL, Alexander CM, Nathanson BH, Haidar T, Dubois RW. Economic and clinical impact of inpatient diabetic hypoglycemia. Endocr Pract. 2009;15:302–12. 10.4158/EP08343.OR

2. Goto A, Arah OA, Goto M, Terauchi Y, Noda M. Severe hypoglycaemia and cardiovascular disease: systematic review and meta-analysis with bias analysis. BMJ. British Medical Journal Publishing Group; 2013;347:f4533. 10.1136/bmj.f4533

3. Yaffe K, Falvey CM, Hamilton N, Harris TB, Simonsick EM, Strotmeyer ES, et al. Association Between Hypoglycemia and Dementia in a Biracial Cohort of Older Adults With Diabetes Mellitus. JAMA Internal Medicine. 2013;173:1300–6. 10.1001/jamainternmed.2013.6176

4. Huang L, Zhu M, Ji J. Association between hypoglycemia and dementia in patients with diabetes: a systematic review and meta-analysis of 1.4 million patients. Diabetology & Metabolic Syndrome. 2022;14:31. 10.1186/s13098-022-00799-9

5. Signorovitch JE, Macaulay D, Diener M, Yan Y, Wu EQ, Gruenberger J-B, et al. Hypoglycaemia and accident risk in people with type 2 diabetes mellitus treated with non-insulin antidiabetes drugs. Diabetes, Obesity and Metabolism. 2013;15:335–41. 10.1111/dom.12031

6. Johnston SS, Conner C, Aagren M, Ruiz K, Bouchard J. Association between hypoglycaemic events and fall-related fractures in Medicare-covered patients with type 2 diabetes. Diabetes, Obesity and Metabolism. 2012;14:634–43. 10.1111/j.1463-1326.2012.01583.x

7. Pratiwi C, Mokoagow MI, Made Kshanti IA, Soewondo P. The risk factors of inpatient hypoglycemia: A systematic review. Heliyon. 2020;6:e03913. 10.1016/j.heliyon.2020.e03913

8. Hashimoto T, Morita A, Hashimoto Y, Yagami F, Sakamoto K, Owada M, et al. Seasonal Variation of Severe Hypoglycemia in Hospitalized Patients 60 years of Age or Older Presenting to an Emergency Center Hospital between 2004 and 2010. Intern Med. The Japanese Society of Internal Medicine; 2013;52:2721–6. 10.2169/internalmedicine.52.0495

9. Minamoto-Higashioka M, Kawamura R, Umakoshi H, Yokomoto-Umakoshi M, Utsunomiya D, Osawa H, et al. Seasonal Variation in Severe Glucose-lowering Drug-induced Hypoglycemia in Patients with Type 2 Diabetes. Intern Med. The Japanese Society of Internal Medicine; 2019;58:1067–72. 10.2169/internalmedicine.1360-18

10. Aspinall SL, Good CB, Jiang R, McCarren M, Dong D, Cunningham FE. Severe Dysglycemia with the Fluoroquinolones: A Class Effect? Clinical Infectious Diseases. 2009;49:402–8. 10.1086/600294

11. Liao S-H, Hu S-Y, How C-K, Hsieh VC-R, Chan C-M, Chiu C-S, et al. Risk for hypoglycemic emergency with levofloxacin use, a population-based propensity score matched nested case-control study. PLOS ONE. Public Library of Science; 2022;17:e0266471. 10.1371/journal.pone.0266471

12. Derijks HJ, Heerdink ER, De Koning FH, Janknegt R, Klungel OH, Egberts AC. The association between antidepressant use and hypoglycaemia in diabetic patients: a nested case–control study. Pharmacoepidemiology and Drug Safety. 2008;17:336–44. 10.1002/pds.1562

13. Tsujimoto T, Yamamoto-Honda R, Kajio H, Kishimoto M, Noto H, Hachiya R, et al. Seasonal Variations of Severe Hypoglycemia in Patients With Type 1 Diabetes Mellitus, Type 2 Diabetes Mellitus, and Non-diabetes Mellitus: Clinical Analysis of 578 Hypoglycemia Cases. Medicine. 2014;93:e148. 10.1097/MD.0000000000000148

14. The National Health and Nutrition Survey Japan, 2019 [Internet]. 2020 [cited 2023 Nov 15]. https://www.mhlw.go.jp/stf/newpage_14156.html. Accessed 15 Nov 2023

15. Ikeda Y, Kubo T, Oda E, Abe M, Tokita S. Incidence rate and patient characteristics of severe hypoglycemia in treated type 2 diabetes mellitus patients in Japan: Retrospective Diagnosis Procedure Combination database analysis. Journal of Diabetes Investigation. 2018;9:925–36. 10.1111/jdi.12778

16. Bruderer SG, Bodmer M, Jick SS, Bader G, Schlienger RG, Meier CR. Incidence of and risk factors for severe hypoglycaemia in treated type 2 diabetes mellitus patients in the UK – a nested case–control analysis. Diabetes, Obesity and Metabolism. 2014;16:801–11. 10.1111/dom.12282

17. Maeda N, Tamagawa T, Niki I, Miura H, Ozawa K, Watanabe G, et al. Increase in insulin release from rat pancreatic islets by quinolone antibiotics. British Journal of Pharmacology. 1996;117:372–6. 10.1111/j.1476-5381.1996.tb15201.x

18. Saraya A, Yokokura M, Gonoi T, Seino S. Effects of fluoroquinolones on insulin secretion and β-cell ATP-sensitive K+ channels. European Journal of Pharmacology. 2004;497:111–7. 10.1016/j.ejphar.2004.06.032

19. Ochi S, Abe M, Shimizu H, Iga J, Ueno S. Hypoglycemia with atypical antipsychotics, but not with typical antipsychotics: A case report. Clinical Neuropsychopharmacology and Therapeutics. 2020;11:5–8. 10.5234/cnpt.11.5

20. Nagamine T. Hypoglycemia associated with insulin hypersecretion following the addition of olanzapine to conventional antipsychotics. Neuropsychiatr Dis Treat. 2006;2:583–5.

21. Suzuki Y, Watanabe J, Fukui N, Ozdemir V, Someya T. Hypoglycaemia induced by second generation antipsychotic agents in schizophrenic non-diabetic patients. BMJ. British Medical Journal

